# Automated diagnosis of usual interstitial pneumonia on chest CT via the mean curvature of isophotes

**DOI:** 10.1101/2025.02.28.25322740

**Authors:** Peter Savadjiev, Morteza Rezanejad, Sahir Bhatnagar, David Camirand, Claude Kauffmann, Kaleem Siddiqi, Ronald J Dandurand, Patrick Bourgouin, Carl Chartrand-Lefebvre, Alexandre Semionov

**Affiliations:** School of Computer Science, McGill University, Montreal, QC, Canada; Centre for Intelligent Machines, McGill University, Montreal, QC, Canada; Rosalind & Morris Goodman Cancer InsCtute, McGill University, Montreal, QC, Canada; Department of Epidemiology, BiostaCsCcs and OccupaConal Health, McGill University, Montreal, QC, Canada; Prime Sciences, Montreal, QC, Canada; Department of Radiology, RadiaCon Oncology and Nuclear Medicine, Université de Montréal, Montreal, QC, Canada; Department of Radiology, Centre Hospitalier de l’Université de Montréal (CHUM), Montreal, QC, Canada; Centre d’intégraCon et d’analyse des données médicales (CITADEL), Centre Hospitalier de l’Université de Montréal (CHUM), Montreal, QC, Canada; Department of Medicine, McGill University, Montreal, QC, Canada; Research InsCtute, McGill University Health Center, Montreal, QC, Canada; Montreal Chest InsCtute, McGill University Health Center, Montreal, QC, Canada; Lakeshore General Hospital, Pointe-Claire, QC, Canada; Department of DiagnosCc Radiology, McGill University, Montreal, QC, Canada

## Abstract

**Purpose:** To test whether the mean curvature of isophotes (MCI), a geometric image transformation, can be used to improve automatic detection on chest CT of Usual Interstitial Pneumonia (UIP), a determining radiological pattern in the diagnosis of Interstitial Lung Diseases (ILD).

**Materials and methods:** This retrospective study included chest CT scans from 234 patients (123 female,111 male; mean age: 61.6 years; age range: 18-90 years) obtained at two independent institutions between 2007 and 2024.

Three different classification models were trained on the original CT images and separately on MCI-transformed CT images: (1) a previously published deep learning model for classifying fibrotic lung disease on chest CT, (2) a classification pipeline based on the EfficientNet-V2 convolutional neural network architecture, and (3) a non-deep-learning model based on the functional principal component analysis (FPCA) of density functions of voxel intensity.

All models were trained on data from the first institution and evaluated on data from the second institution with the recall-macro, precision-macro and F1-macro scores. Performance difference between classifier pairs was tested with the Stuart-Maxwell marginal homogeneity test.

**Results:** For a fixed model architecture and training algorithm, MCI-transformed images yield comparable or better classification performance than the original CT images. The best performance improvement achieved with MCI compared to CT was: recall-macro 0.83 vs 0.57, precision-macro 0.81 vs 0.50, F1-macro 0.80 vs 0.49, p=4.2e-5.

**Conclusion:** MCI may be a valuable addition to existing AI systems for screening for UIP on chest CT.

**Summary statement:** Machine learning methods for identifying usual interstitial pneumonia on chest CT perform better when the input CT images are transformed via the mean curvature of isophotes (MCI), a geometric transformation method known from classical computer vision.

**Key Points:**

- Three machine learning models were trained on a dataset of 158 patients from one institution and tested on another dataset of 76 patients from an independent institution to discriminate for usual interstitial pneumonia (UIP) on chest CT in a 3-group classification task.
- When keeping the network architecture and parameters fixed, changing the input image domain from the original CT to MCI-transformed images improved classification performance (Stuart-Maxwell test, p < 5e-3)
- MCI may be a valuable addition to existing machine learning systems for screening for UIP on chest CT, whether based on deep learning or on simpler shallow classifiers.

## Introduction

Interstitial lung disease (ILD) is a heterogeneous group of disorders characterized by architectural changes of the lung parenchyma, resulting from progressive pulmonary inflammation and/or fibrosis. These changes lead to deterioration of lung function, disability, and premature death [1].

The standard diagnostic test for ILD is a high-resolution computed tomography (CT) scan of the chest [2]. Unfortunately, CT interpretative guidelines for ILD are largely subjective, and radiological diagnosis of ILD suffers from important inter-reader variability [2]. As a result, much effort has gone into developing quantitative CT analysis algorithms usually based on a deep-learning strategy [2]. Due to their success in a wide range of applications, it is commonly believed that deep learning algorithms such as convolutional neural networks (CNNs) extract the most effective image features for a given task, whereas human-engineered methods derived from domain knowledge are assumed to be sub-optimal.

However, a growing body of literature shows that in fact, CNN performance can be significantly improved if combined with explicit domain knowledge [3–6]. In particular, the mean curvature of isophotes (MCI) is a geometric image transformation that has recently been shown to improve CNN performance in detecting chronic obstructive pulmonary disease (COPD) on chest CT [7,8].

In the present work, we take the MCI framework to the domain of ILD. At the heart of our analysis lie two CNN-based methods. First is the SOFIA method, a well-known and well-cited approach to classifying fibrotic lung disease on chest CT [9,10]. Since the SOFIA method is older and does not use a contemporary CNN architecture, we also implemented a second analysis using a recent EfficientNet-V2 CNN architecture [11]. Our main hypothesis is that regardless of the specific CNN architecture choice, changing the input image domain from the original CT to MCI-transformed images, while keeping the network architecture and parameters fixed, can improve performance in a 3-group classification task.

These 3 groups consist of control CT scans without pulmonary disease; CT scans with ILD featuring a particular radiological pattern known as usual interstitial pneumonia (UIP) [12,13] ; and CT scans with other types of lung disease including types of ILD other than UIP. We stratify the ILD cases on a UIP vs. non-UIP basis because the UIP pattern has been associated with a faster progression and worse clinical prognosis. In this manner we emulate a recent clinical trial on progressive fibrosing ILD, where randomization was stratified according to the presence of UIP vs other ILD patterns [1]. As a result of this trial and others, anti-fibrotic therapy is now available following an UIP diagnosis.

Our secondary hypothesis is that the mathematical properties of the MCI transformation will make it possible for a simpler computational alternative based on functional principal components analysis (FPCA) [14,15] to yield a classification performance comparable to that of deep learning.

We worked with imaging data from two independent institutions, with data from one institution used to train classification models and data from the second institution used for independent testing.

## Materials and Methods

### Patient populations

This retrospective study was conducted with data acquired between 2007 and 2021 at McGill University and its affiliated hospitals, Montréal, QC, Canada (Institution 1), and between 2015 and 2024 at the Centre Hospitalier de l’Université de Montréal (CHUM), Montréal, QC, Canada (Institution 2). The study received ethics approval at each participating institution, and patient informed consent was waived.

In both institutions, ILD cases are initially diagnosed during multidisciplinary meetings between pulmonologists, rheumatologists, pathologists and radiologists. Cases included in this study were randomly selected from such interdisciplinary discussions. Scans with non-ILD pulmonary diseases were randomly selected from participating thoracic radiologists’ daily worklist using search keywords: sarcoidosis, Kaposi sarcoma, bronchioloalveolar carcinoma, pneumonia, lymphangitic carcinomatosis, pulmonary edema, pulmonary alveolar proteinosis, atelectasis, PCP, and COVID19. The final diagnosis based on chest CT and available clinical history for each case was done by thoracic radiologists: AS (14y experience) at Institution 1; PB (5y experience), and CCL (26y experience) at Institution 2. Cases with avert pulmonary parenchymal and pleural disease which had no inkling of possible ILD were excluded; for example, cases with large pleural effusions, extensive atelectasis, multilobar pneumonia, lobectomy/pneumonectomy, or diffuse nodular metastases.

Scans of patients without radiological evidence of pulmonary disease other than occasional micronodules were selected as normal controls. Nine such scans were initially retrieved in Institution 1. To increase the number of controls in the training dataset, 27 additional control scans were selected from the clinical practice of a participating respirologist (RJD, 44y experience) affiliated with Institution 1. These 27 patients are a subset of the control patient cohort previously reported in [7].

DICOM images were extracted from the radiology databases (PACS) of the two institutions. All CT images were anonymized prior to further analysis.

### Diagnostic groups

The official 2018 clinical practice guideline for radiological diagnosis of ILD [12] advocates the use of four diagnostic categories: UIP, Probable UIP, Indeterminate for UIP, Alternative Diagnosis. In an update from 2022 [13], the guideline committee discusses the option of merging the ‘UIP’ and ‘Probable UIP’ categories, and provides several reasons for doing so. In our experiments, we merged the ‘UIP’ and ‘probable UIP’ groups into one. We also merged the ‘Indeterminate for UIP’ group, the ‘Alternative Diagnosis’ group as well as the non-ILD chronic pulmonary disease patients into another group. In this manner, a final set of three diagnostic groups was obtained: (1) patients with UIP or probable UIP, (2) patients with chronic pulmonary disease without evidence for UIP (may or may not be part of the ILD spectrum), and (3) the control patients without pulmonary disease.

### CT Imaging characteristics

Each of the two institutions represents a large academic network of participating hospitals, thus patient scans were acquired at different sites with different scanner models and parameters, as detailed in Table 1.

**Table 1.**
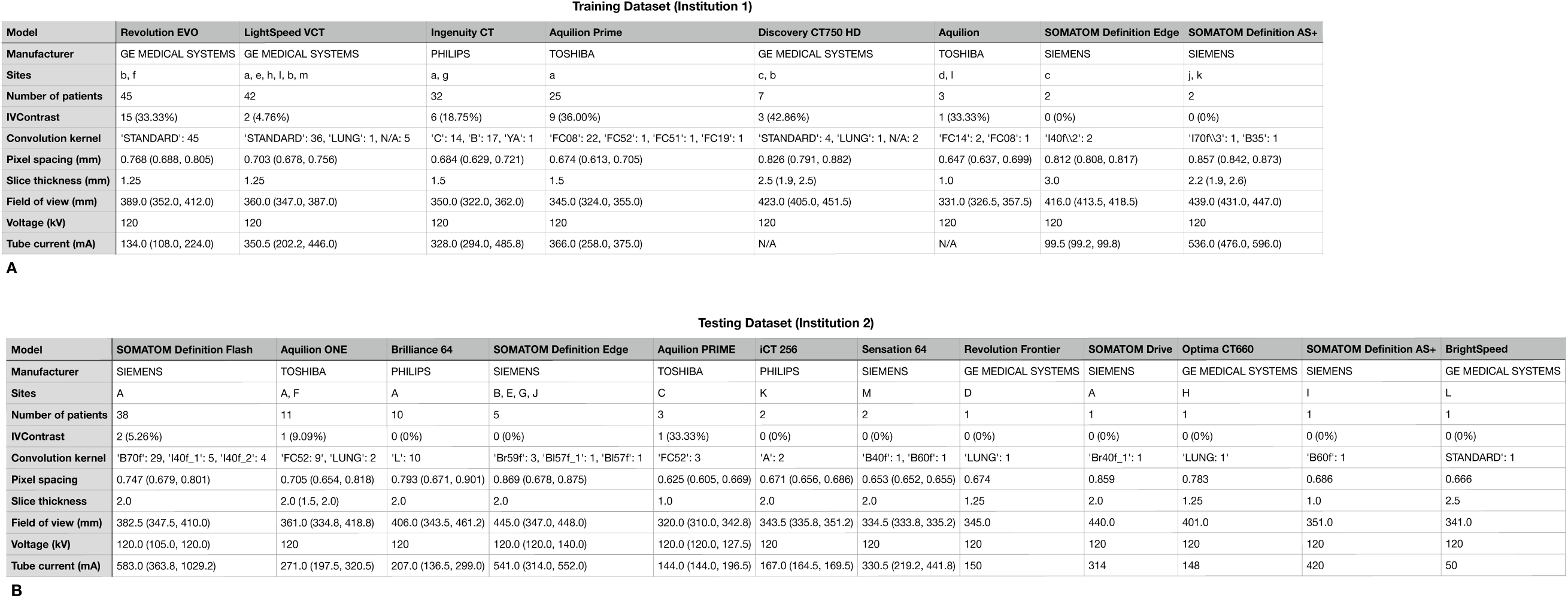
Scan parameters for the training (A) and testing (B) datasets. For each scanner model used, the sites at which this model was used are identified with lower-case letters for the training dataset (A) and capital letters for the testing dataset (B). The table also shows the number of patients scanned with this scanner model, the number among them that was scanned with intravenous contrast (IVContrast, data shown as a number and also as a percentage of the total number of patients for that scanner), the convolution kernel used and the number of scans corresponding to that kernel, as well as several quantitative parameters. If a given quantitative parameter has a single value for all patients for that scanner model, that value is given. If more than one value is present within that patient group, the median (interquartile range) is provided. N/A: data not available

### SOFIA

The Systematic Objective Fibrotic Imaging Analysis (SOFIA) algorithm was introduced in [9] as a chest CT classification method based on the Inception-ResNet-v2 CNN architecture. A particular characteristic of SOFIA is the data augmentation technique, which consists in dividing the craniocaudal lung length into 4 equal blocks of segmented axial images, then selecting at random one slice from each block and joining each selected slice into a 2x2 slice montage, as shown in Fig. 2 in [9]. This process is repeated 500 times for each 3D scan, to create 500 unique random montages per scan. Each montage was downsized to 350x350 pixels. The Inception-ResNet-v2 architecture was then trained on the resulting montages. See [9] for details.

### Three image contrasts

To assess the benefit of the MCI transformation, we compared the diagnostic performance of the SOFIA approach trained on CT images to its performance when trained separately on MCI images.

To create the SOFIA montages, we first selected random slices from the CT images. Then, the same slice selections were used to create montages from the corresponding MCI images. In this manner, the set of montages differed only in image contrast (CT vs MCI), not in slice selection.

While working with the SOFIA method, we hypothesized that when 4 slices are downsized and combined together into a single 350x350 montage, the main discriminative signal carried by each montage would reflect mostly the overall shape of the slices, as opposed to smaller scale textural features. This hypothesis was also motivated by theoretical considerations and previous findings reviewed in the Discussion section.

To test this hypothesis, we created a third set of montages by binarizing the CT montages, so that all voxels within the lung mask were set to 1 (in all three montage sets, voxels in the background were set to 0).

Fig 1. shows an example montage for each of the three types of images.

**Figure. 1.**
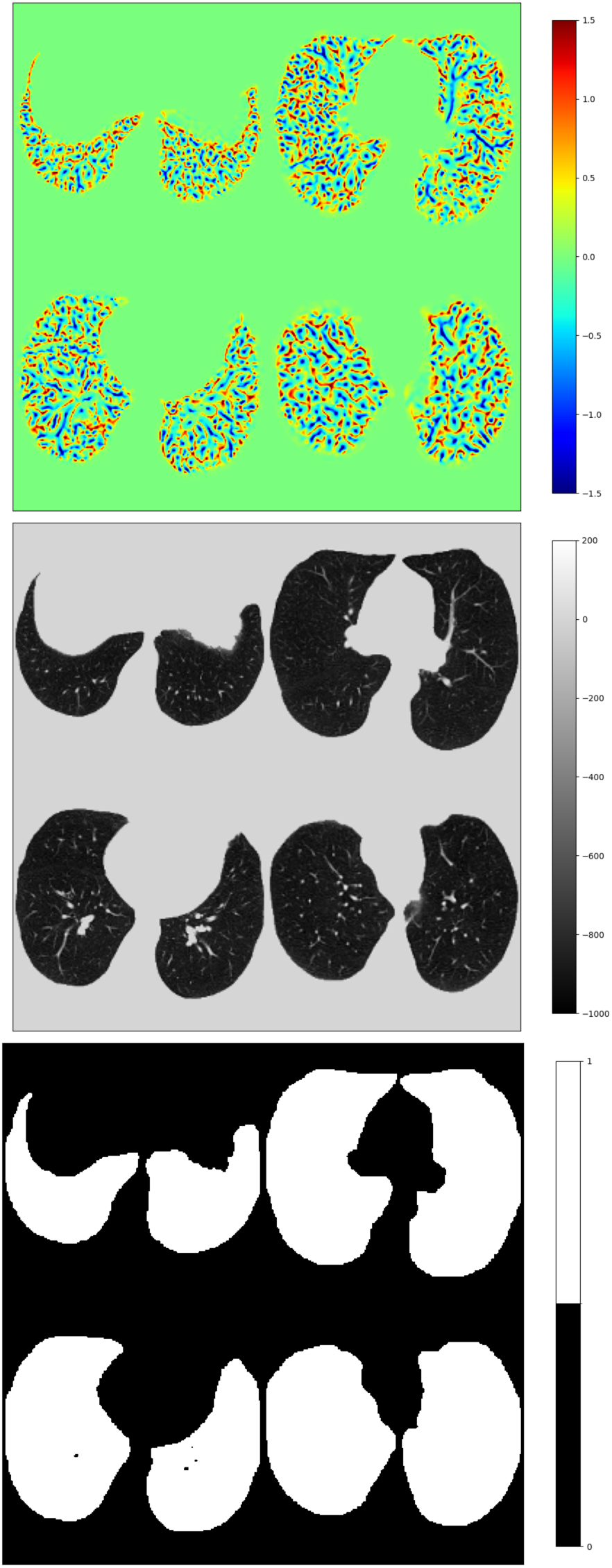
Examples of MCI (top), CT (middle) and binary shape (bottom) 2x2 montages used as input to the SOFIA method.

### Image Classification with EfficientNetV2

The SOFIA method was published in 2018 [9] and uses the Inception-ResNet-V2 architecture originally published in 2017 [16]. Recognizing the advances in deep learning made since then, we also implement a classification pipeline based on a contemporary CNN architecture: EfficientNet-V2 [11]. As the choice and design of a 2x2 montage as used in SOFIA appears somewhat arbitrary, we created a simplified set of inputs to the EfficientNet-V2 architecture, by taking all individual axial slices of a scan except the top and bottom 10%, and used them as input on their own, without creating multiple slice montages. Fig. 2 shows examples of such single slice inputs.

**Figure. 2.**
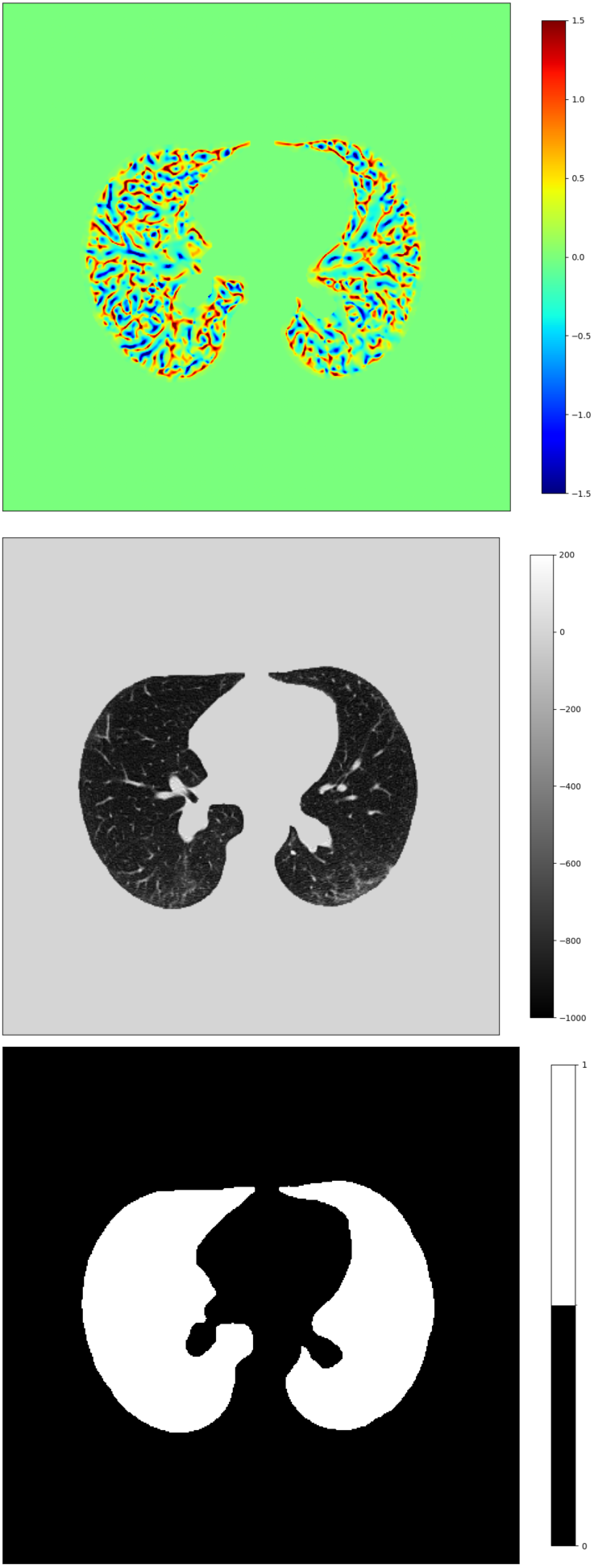
Examples of MCI (top), CT (middle) and binary shape (bottom) single slice inputs used as input to our Simplified SOFIA method.

More details on the implementation of the SOFIA and EfficientNet-V2 pipelines are provided in the Supplementary Material.

### Alternative classification based on FPCA of voxel distributions

We also compared the performance of SOFIA to that of a simpler method based on the FPCA of voxel intensity distributions [14,15]. Details are provided in the Supplementary Material.

### Model evaluation

We report the classification performance of all our models in terms of three standard machine learning metrics: precision, recall and F1-score. Since there are more than 2 classes, we report the per-class macro average of these metrics.

More specifically, in a binary classifier, specificity is equivalent to the sensitivity of the negative class. However, the concepts of ‘positive’ and ‘negative’ class are not defined when there are more than two classes, as in our experimental setting. In a binary setting, the recall measure is equivalent to sensitivity. In a multi-class setting, the per-class macro average of recall is then equivalent to balanced accuracy, i.e., the average of sensitivity computed for each individual class. In the special case of a binary classification, balanced accuracy is simply the average of the sensitivity and specificity. As for the precision-macro value, it is the multiclass equivalent of the average of the positive and negative predictive values of a binary classifier.

### Statistics

To compare classifier pairs, we apply the Stuart-Maxwell marginal homogeneity test [17,18]. It is a generalization of the McNemar test to the case of more than two categories (three in our case). We performed 5 comparisons, which implies that following a Bonferroni correction for multiple comparisons, the p-value threshold for significance is 0.05/5=0.01.

### Software

The computation of MCI was done in Matlab with code provided publicly by [7]. While code for the Inception-ResNet-v2 model itself is publicly available, the overall SOFIA pipeline is not, so we reimplemented SOFIA based on details available in [9]. CNN training and evaluation was done in Python 3.10.12 with PyTorch 2.1.2+cu121. In particular, we used the CNN implementations available in the timm python package v.0.9.2 with model identifiers ‘inception_resnet_v2’ and ‘tf_efficientnetv2_s’. FPCA modeling was done with a combination of R,Python and Matlab programming languages, based on code provided at [https://github.com/czhang-pstat/Density-Review]. The Stuart-Maxwell test was computed with the homogeneity() function from the statsmodels python package v.0.14.4. Our code is provided at the following link: https://github.com/petersv2/MCI_UIP/

## Results

The final training dataset (institution 1) consisted of 158 patients, and the final testing dataset (institution 2) comprised 76 patients.

Table 2 presents the patient population within each dataset. The two datasets are age-matched (p=0.19, Mann-Whitney U test).

**Table 2.** Mean age [range], Median age [interquartile range], total number of patients (number of female patients), and number of patients in each diagnostic group for each dataset.

|  | Institution 1 (training) | Institution 2 (testing) |
| --- | --- | --- |
| Age at scan in years,<br>mean [range] | 62.1 [19.0, 86.0] | 60.5 [18.0, 90.0] |
| Age at scan in years,<br>median [interquartile range] | 65.0 [53.0, 72.8] | 62.0 [54.0, 68.0] |
| Total number of patients<br>(number of females) | 158 (81) | 76 (42) |
| Group 1 (controls) | 36 | 16 |
| Group 2<br>(non-UIP lung disease) | 74 | 48 |
| Group 3<br>(probable or definite UIP) | 48 | 12 |

Table 3 presents the diagnostic performance of the classification models on the testing dataset from Institution 2.

**Table 3.** Diagnostic performance of classification models.

| Method | Image Contrast | Classifier Performance metrics |  |  |
| --- | --- | --- | --- | --- |
|  |  | Recall-macro<br>(balanced accuracy) | Precision-macro | F1-macro |
| **SOFIA** | MCI | 0.72 | 0.62 | 0.65 |
|  | CT | 0.65 | 0.62 | 0.63 |
|  | Binary/Shape | 0.56 | 0.53 | 0.54 |
| **EfficientNet-V2** | MCI | 0.74 | 0.80 | 0.75 |
|  | CT | 0.72 | 0.67 | 0.67 |
|  | Binary/Shape | 0.63 | 0.58 | 0.60 |
| **FPCA** | MCI | 0.83 | 0.81 | 0.80 |
|  | CT | 0.57 | 0.50 | 0.49 |

Table 4 shows the p-value of the Stuart-Maxwell test comparing MCI vs CT classifiers in the three settings under investigation, as well as the improvement in MCI classification performance going from the original SOFIA to the simplified SOFIA and then to the FPCA framework.

**Table 4.** P-values of the Stuart-Maxwell marginal homogeneity test, for selected pairs of classifiers.

| Method | Image Contrast | Stuart-Maxwell test p-value |
| --- | --- | --- |
| SOFIA | MCI | 0.072 |
|  | CT |  |
| EfficientNet-V2 | MCI | 4.7E-03 |
|  | CT |  |
| FPCA | MCI | 4.2E-05 |
|  | CT |  |
| SOFIA | MCI | 3.4E-04 |
| EfficientNet-V2 |  |  |
| EfficientNet-V2 | MCI | 4.8E-04 |
| FPCA |  |  |

## Discussion

MCI is an image transformation which reflects the geometric structure of the imaged scene. It has long been studied in the field of computer vision and other fields such as astronomy due to its ability to extract the intrinsic structure of images in an invariant manner [7,8,19].

It is widely assumed that CNNs automatically discover the optimal features for a task. However, in chest CT analysis, it was shown that a CNN performs better at detecting early COPD when presented with MCI-transformed images, as opposed to the original CT images [7]. This is not because MCI images contain more information - since they are derived from CT images, all information in MCI is necessarily present also in the CT contrast. However, the invariance properties of MCI reduce unwanted variability while at the same time reveal geometric information that appears helpful in detecting/characterizing lung disease, information which the CNN did not seem to recover on its own from the original CT [7].

In this study, we took this line of work to the field of ILD, with the task of differentiating three classes of images: (1) controls without pulmonary disease , (2) patients with probable or definite UIP, and (3) patients with other pulmonary disease, including (but not limited to) patients with non-UIP ILD. In this manner, we address a challenging classification problem of pulmonary diseases not limited to the ILD spectrum. This is in contrast to a number of studies which focus only on binary UIP vs non-UIP classification [20–23]. While some train their models on populations that include non-ILD pulmonary disease and healthy controls [22,23], testing is usually done on ILD-only cohorts.

In addition to diagnostic heterogeneity, our data is also highly heterogeneous in terms of imaging characteristics. As seen in Table 1, the training data contains images acquired at 13 scanning sites, with 8 different scanner models and a large variability of scanning parameters, including 23% of scans (36/158) with intravenous contrast. The testing data was acquired at 13 sites different from those of the training data, on 12 scanner models, with 5% of scans (4/76) having intravenous contrast. In this manner, we attempted to create minimally-curated datasets representative of everyday clinical practice.

In the context of such data heterogeneity, the objective was to showcase the relative improvement in performance for the same model architecture and training algorithm when provided MCI-transformed images as input, as opposed to original CT images. Model training and testing was done on independent datasets from two institutions.

Our first comparison between MCI and CT was done through SOFIA [9,10], a method using the Inception-ResNet-v2 CNN architecture [16]. While Inception-ResNet-v2 is an older architecture, we chose to work with SOFIA as it is a well-known and well-cited approach to CT image analysis in the ILD literature. We next tested a classification pipeline based on the more contemporary EfficientNet-V2 architecture [11]. Finally, we tested a non-deep-learning classifier based on the FPCA of density functions of voxel intensity [14,15].

Our results in Tables 3 and 4 show that for a fixed CNN architecture, MCI gives comparable or better results than CT. With the older SOFIA approach, MCI performance is comparable to that of CT (balanced accuracy of 0.72 vs 0.65, precision-macro of 0.62 vs 0.62, F1-macro of 0.65 vs 0.63, p=0.072). With the more recent EfficientNet-V2 approach, both MCI and CT contrasts produce improved classification relative to SOFIA results. Furthermore, with EfficientNet-V2, the observed improvement in MCI classification relative to CT classification is now significant. (balanced accuracy of 0.74 vs 0.72, precision-macro of 0.80 vs 0.67, F1-macro of 0.75 vs 0.67, p=4.7e-3). The best performance is obtained with MCI using the simpler FPCA technique (balanced accuracy of 0.83 vs 0.57, precision-macro of 0.81 vs 0.50, p=4.2e-5).

It may be argued that the CNN could have achieved better performance with CT images, without requiring explicit MCI information, by optimizing the training procedure and hyperparameter choices, or with better data-augmentation. This may well be the case; however, we restate that the objective of this study was not to determine the most optimal CNN model for this specific clinical problem. Rather, it was to showcase the relative performance improvement for the same CNN method when provided MCI images as input, as opposed to original CT images.

A recent deep-learning study for detecting UIP on chest CT claimed that the low prevalence of UIP will always result in a low positive predictive value (PPV) for any tool developed to screen for or classify ILD in a healthcare-based cohort [23]. In that binary classification study, their deep learning model resulted in a PPV of 68% and negative predictive value (NPV) of 87% [23]. In our study, MCI achieved a precision-macro value of 80% with EfficientNet-V2 and 81% with FPCA. As a reminder, the precision-macro value is the multiclass equivalent of the average of the PPV and NPV of a binary classifier. In [23], the average of PPV and NPV was 78%.

Our results also link to an ongoing debate on whether CNNs focus preferentially on texture or on shape [24]. In our study the CNN has a clear preference for shape. In a 3-class task, a random classifier is expected to result in a balanced accuracy of 0.33 purely by chance. Here, we observed that the binary (textureless) shape images achieve a much higher value, between 0.56 and 0.63, which nearly double the value expected by pure chance and only 10% (or less) below the performance achieved with the CT contrast. One implication is that lung shape alone plays a role in distinguishing between the 3 groups of patients. Another implication is that clearly, CNNs over-rely on shape and under-exploit CT texture. It appears that due to shortcut learning [25,26], the CNN learns the simplest possible signal that leads to discrimination (also known as simplicity bias), which is this case is carried mostly by the shape of the segmented lung slices over a uniform background. Again, this runs contrary to the commonly held expectation that CNNs find on their own the best features for a given task, as overall lung shape is clearly not the best way to detect UIP. Our work shows that the same CNN can achieve better results if explicitly provided with precomputed MCI information. Interestingly, even better performance is achieved by the simpler FPCA method which by design does not use shape information.

The main limitation of our study is the small sized datasets. As this is an exploratory study, we strived to present a proof-of-concept of the applicability of MCI in ILD in the presence of data heterogeneity and with testing on an independent external dataset. In future work, we will expand testing with much larger datasets such as those collected by the Open Source Imaging Consortium [27][https://www.osicild.org/]. Another limitation is that we did not examine associations between classification results and clinical outcomes. As a proof-of-concept, our study focused on radiological data only. In future work will also focus on clinical outcomes with larger datasets as described above. Finally, one technical limitation is that we only worked with MCI at a single spatial scale of 2mm. As discussed in [7], MCI is derived from classical scale-space literature in computer vision, where scale is modeled as a continuously varying parameter. In future work, we will extend our approach to a range of spatial scales with an optimal scale being determined automatically akin to classical scale-space methods in computer vision. [28,29] Whereas this study showcased the utility of MCI for discrimination, an important next step is understanding the association between the MCI signal and underlying biological tissue characteristics. A recent histology study on explanted lungs with radiologically documented interstitial lung abnormalities found varying degrees of fibrosis in areas that appeared normal on CT scans [30]. An interesting topic of future investigation is whether MCI can reveal abnormal structure in such normal-appearing areas on CT. Future work will also focus on the role MCI might play in prediction of disease progression and response to treatment [2,27], as well as in the building of foundation models for lung imaging [31].

In conclusion, our results suggest MCI may be a valuable addition to existing machine learning systems for screening for UIP on chest CT, whether based on deep learning or on simpler shallow classifiers. In this work, we transform the input image space with MCI, but it is worthwhile exploring whether a MCI computation layer can be incorporated directly within a deep learning architecture.

## Data Availability

The images and data used in this study are hospital patient records, containing protected healthcare information (PHI). As such, we do not have the authority to share them or make them public.

## Acknowledgements

PS acknowledges funding from NSERC Discovery Grant RGPIN-2018-05636. SB acknowledges funding from NSERC Discovery Grant RGPIN-2020-05133. PS is currently employed at the Department of Radiology, University of Vermont, Burlington VT USA.

## Supplementary Material

Note: a list of references specific to the supplementary material appears at the end of this document

### Supplementary methods

#### Image preprocessing and MCI computation

Following the methodology of Savadjiev et al. [1], we first resampled all original CT images to an isotropic voxel grid of 0.9×0.9×0.9 mm. The MCI transformation of these resampled images was then computed, with a scale value of σ=2mm [1]. A lung mask was computed on the resampled CT images using the Chest Imaging Platform (http://chestimagingplatform.org/), an open-source software. This mask was then applied to both the CT and the MCI-transformed images to set voxels outside the lungs to 0.

#### SOFIA pipeline

We followed all the details provided by the original SOFIA publication [2] to reimplement their algorithmic pipeline. As described in [2] and as visualized in Figure 1 in this paper, 500 random 2x2 axial slice montages were created from the chest CT scan of each patient, and were then used as input to the Inception-ResNet-v2 CNN architecture. A total of 79,000 such montages were created from the training dataset and 38,000 from the testing dataset.

The Inception-RestNet-V2 model we used is the one supplied by the timm python library v.0.9.2, under the identifier ‘inception_resnet_v2’: https://huggingface.co/timm/inception_resnet_v2.tf_in1k

Replicating the approach described in [2], we used this model without pre-training.

#### EfficientNet-V2 pipeline

Instead of creating slice montages, we simply cropped the masked axial slice around its tight-fitting bounding box and then added the necessary padding of 0-valued pixels around it to obtain a final image with size 384x384 pixels, without additional interpolation/resizing. As a result, our training dataset comprised 34841 images of size 384x384, and the testing dataset comprised 17425 images of the same size. No further data augmentation or hyper parameter optimization was performed. We used default parameters for the learning rate optimizer and scheduler in PyTorch.

The EfficientNet-V2 model we used is the one supplied by the timm python library v.0.9.2, under the identifier ‘tf_efficientnetv2_s’: https://huggingface.co/timm/tf_efficientnetv2_s.in21k_ft_in1k

This is a relatively small model with 21.5 M parameters. We used this model with pre-training, which has first been done on ImageNet-21k and then on ImageNet-1k. In our pipeline this pretrained model was then fine-tuned on our own training dataset.

#### Per-patient classification

The CNN classification approach for SOFIA (or EfficientNet-V2) was applied separately on the 3 sets of montages (or individual slices, respectively) described above and in the main manuscript. In other words, the three different image contrasts were not used as three channels in the same model. Rather, three different models were trained for each individual image contrast.

At inference time, each of these trained models assigns a classification label to each montage (or individual slice, respectively). To obtain a per-patient label, we used majority voting, i.e., the final predicted label assigned to each patient was the label associated with the largest number of montages (slices, respectively) pertaining to that patient.

#### Alternative classification based on FPCA of voxel distributions

Inspired by the LAA950 metric commonly used in lung quantitative CT analyses [3], the work by Savadjiev et al. [1] introduced a metric called “hmci1” defined as the fraction of lung voxels with an MCI value larger than a certain threshold. Equivalently, this can be seen as the area under the probability density function of MCI values above a given threshold. In [1] an MCI threshold value of 1mm^-1^ was chosen somewhat arbitrarily.

Recognizing the limitations of fixing a specific threshold value, here we work with the entire MCI probability density function as a data object itself, obviating the need for choosing a threshold. This is done within a functional principal components analysis (FPCA) framework, where low-dimensional embeddings of the density functions are obtained by computing the principal components (modes of variation) of the MCI density functions [4,5]. These embeddings are then used for classification.

Probability densities (PD) are nonnegative functions with an integral that sums to 1. These constraints imply that the functional space of PDs is convex but not linear. As a result, standard FPCA methods are not directly applicable. To address this, Petersen et al. [4] have proposed a transformation approach where PDs are mapped to a Hilbert space of functions through a continuous and invertible map. The resulting representations of PDs in this Hilbert space are then amenable to the construction of functional modes of variation and FPCA. Results are mapped back to the density space using the inverse transformation map.

In this work, we used the implementation of this method available at [https://github.com/czhang-pstat/Density-Review] to build a simple classification model based on the modes of variation of the MCI densities. We computed the PDs of MCI values over the extent of the segmented lungs, using kernel density estimation (function ksdensity() in Matlab). We then applied the method of [4] with their log quantile density (LQD) transformation to compute FPCA embeddings of all the MCI PDs from the training dataset. The MCI PDs from the testing dataset were transformed in the same fashion and projected into the FPCA space computed from the training PDs. This was achieved with the MdVar() function in [https://github.com/czhang-pstat/Density-Review]. Classification of the FPCA embeddings of the testing PDs was achieved using a k-nearest-neighbor classifier (k=5). For comparison, the same process was repeated with the HU values of CT images over the same lung segmentations.

